# Social contact patterns during the COVID-19 pandemic in 21 European countries – evidence from a two-year study

**DOI:** 10.1101/2022.07.25.22277998

**Authors:** Kerry LM Wong, Amy Gimma, Pietro Coletti, CoMix Europe Working Group, Christel Faes, Philippe Beutels, Niel Hens, Veronika K Jaeger, Andre Karch, Helen Johnson, W John Edmunds, Christopher I Jarvis

## Abstract

Most countries have enacted some restrictions to reduce social contacts to slow down disease transmission during the COVID-19 pandemic. For nearly two years, individuals likely also adopted new behaviours to avoid pathogen exposure based on personal circumstances. We aimed to understand the way in which different factors affect social contacts – a critical step to improving future pandemic responses. The analysis was based on repeated cross-sectional contact survey data collected in 21 European countries between March 2020 and March 2022. We calculated the mean daily contacts reported using a clustered bootstrap by country and by settings (at home, at work, or in other settings). Where data were available, contact rates during the study period were compared with rates recorded prior to the pandemic. We fitted censored individual-level generalized additive mixed models to examine the effects of various factors on the number of social contacts. The survey recorded 463,336 observations from 96,456 participants. In all countries where comparison data were available, contact rates over the previous two years were substantially lower than those seen prior to the pandemic (approximately from over 10 to <5), predominantly due to fewer contacts outside the home. Government restrictions imposed immediate effect on contacts, and these effects lingered after the restrictions were lifted. Across countries, the relationships between national policy, individual perceptions, or personal circumstances determining contacts varied. Our study, coordinated at the regional level, provides important insights into the understanding of the factors associated with social contacts to support future infectious disease outbreak responses.

## Introduction

COVID-19 primarily spreads via aerosols and thus through close, direct, and in-person social contacts. Especially at the start of the COVID-19 pandemic, when there were no effective vaccines and treatments, the spread of the disease was mostly managed by promoting non-pharmaceutical interventions (NPIs), which served to avoid potential exposure to the pathogen in the population^1^.

In Europe, Italy was the first country to begin major NPIs. On March 9 2020, the country issued their first national lockdown mandating 60 million residents to stay at home^2^. On March 14, the Spanish government declared a state of emergency^3^. Similar announcement emerged in other European countries soon thereafter^4^. Lockdown mandates posed immediate restrictions on social contacts. Contact surveys conducted quickly after these measures showed that the number of mean daily contacts per person rapidly fell from pre-pandemic levels to approximately 4 per person in Belgium, Italy, Netherlands, Germany, United Kingdom (UK), France, Greece, and Luxembourg^5–10^.

Many countries in the region began to ease restrictions during the summer of 2020 and continued to vary the intensities of various NPIs to control the epidemic in the two years that followed. Although contacts curbed when restrictions were introduced, commensurate rises in social contacts were not seen when measures were lifted. In the UK, for instance, at its highest recorded level, the mean number of contacts in 2021 only returned to approximately 50% of that observed in pre-pandemic times (as measured in the POLYMOD contact survey study in 2005 to 2006^11^)^12^. Social contact behaviours during the pandemic have been shown to vary by population and demographics^12,13^. In addition to the guidance issued by the authorities, individuals may also have adopted new behaviours to avoid pathogen exposure for other reasons, e.g., personal circumstances such as risk profile, perceived pandemic threat level, work requirements, among others^14^. Research opportunities into the complexity and determinants of changes in social contacts have thus far been relatively limited.

We conducted a multi-country study in the European region, following individuals in real-time over the course of the COVID-19 pandemic. We aimed to assess social contacts in 21 European countries between March 2020 and March 2022 using a large dataset. We present descriptive analyses showing the mean number of contacts reported by study participants, and how these changed over time across the study countries. We also examine the potential determinants influencing the mean number of contacts.

## Methods

### Study design

CoMix is an online multi-country behavioural survey conducted in 21 European countries between March 2020 and March 2022. In each study country, a nationally representative sample was recruited using quota sampling based on age, gender, geographic region, and where possible, socioeconomic status to reflect the distribution within the national population. The market research company Ipsos-MORI or a local partner conducted participant recruitment through a combination of social media, web advertising, and email campaigns to meet quotas.

The design of the CoMix survey is based on the POLYMOD contact survey. The POLYMOD survey is a self-administered paper survey in the form of a daily diary recording participants’ social contacts^11^. In the CoMix study, participants consented to self-report their social contacts made on the day prior to survey participation. Other survey questions in CoMix included participants’ household composition, work attendance status, presentation of symptoms common to respiratory illnesses, COVID-19 vaccination (since December 2020), among others. Details of the CoMix study including the protocol, methodology, and survey instrument have been published^15–17^.

### Study participants

CoMix was first launched in March 2020 in the UK, and in April 2020 in Belgium, and Netherlands. Participants were invited to partake up to 10 survey rounds, with each round being two weeks apart. - A panel of approximately 1500 participants aged 18 or above were recruited in each of Belgium and the Netherlands. While most data were collected on behaviours in adults, a proportion of the respondents reported contacts on behalf of their children. In the UK, two concurrent panels of respondents were surveyed in alternating weeks; and since August 2020, the number of participants in each panel increased from approximately 1500 to approximately 3000. Additionally, we included data from the German COVIMOD study, a similar contact survey study first began in April 2020^18^. New participants were recruited on a rolling basis to replenish the sample as participants dropped out of the study in the UK, Belgium, Netherlands and Germany.

From December 2020, 17 other countries were added to the study:

- 7 countries – Austria, Denmark, France, Italy, Poland, Portugal, and Spain – between December 2020 and April 2021 (referred to as Group 1 countries)
- 2 countries – Greece and Slovenia – between February 2021 and June 2021, and 3 countries – Switzerland, Finland, and Lithuania – between February 2021 and October 2021 (referred to as Group 2 countries)
- 4 countries – Hungary, Slovakia, Estonia, and Croatia – between May 2021 and October 2021, and lastly Malta between May 2021 and August 2021 (referred to as Group 3 countries)

In each of these 17 countries, the adult panel (aged 18 or above) comprised of at least 1500 participants that were invited to 7 survey rounds (with the exception of Switzerland, Finland, and Lithuania, where 13 survey rounds were conducted), and another child panel (aged 0-18 years) comprised of at least 300 children who were invited to 2 survey rounds. Where possible, one child survey round was timed to be rolled out during school period, and the other during school closure (closure might have been due to COVID-19 or school holiday). In all countries, parents (at least 18 years old) completed the surveys on behalf of one of their children (<18 years old) who lived in the same household. In this analysis, all 21 countries are collectively referred to as CoMix+ countries.

### Data

#### Reporting of contacts

Contacts that occurred on the day prior to the survey were reported in two ways: individual contacts and group contacts. Participants were asked to list each individual contact and its characteristics separately. Participants also had to option to report the total number of contacts (“group contacts”) they had at home, work, or other settings, both overall and for physical contacts only. Other settings included, for instance, a place of worship, essential and non-essential shops, a place of entertainment such as restaurant, bar, cinema, or a place for sports. We defined direct contact as anyone who met the participant in person with whom at least a few words were exchanged or physical contact was made. Questions on group contacts were included at the end of the survey, and they were added to surveys from May 2020 onwards to accommodate individuals – such as those working in patient- or public-facing roles – who could not record details of all individual contacts that they made. Where possible, we compared the mean number of contacts reported by CoMix participants during the COVID-19 pandemic to that presented in the POLYMOD study^11^.

#### Demographic information

The survey captured information about participants demographics, and whether participants attended work. We grouped adult participants by age into the groups of 18-29, 30-39, 40-49, 50-59, 60-69, and 70 years and above. Participants were asked to report how they describe their gender, with the options of “Female,” “Male,” “In another way,” or “Prefer not to answer.”

#### Risk perception, status, and mitigation

Participants were asked questions about their uptake of risk mitigating activities and asked to respond to statements regarding their perception of risk. Participants were asked to respond to the following statements: (i) “I am likely to catch coronavirus”; (ii) “I am worried that I might spread coronavirus to someone who is vulnerable”; and (iii) “Coronavirus would be a serious illness for me” with the Likert scale of “Strongly agree,” “Tend to agree,” “Neutral”, “Tend to disagree,” and “Strongly disagree”. Participants self-reported whether they considered themselves to be high risk based on definitions given in the survey, which changed between survey versions as government advice changed (see questionnaires for details). Participants were also asked whether they wore a face covering and in which settings, and their COVID-19 vaccination status (since December 2020).

#### Presentation of COVID-like symptoms

Participants were asked to report the presentation of COVID-19-compatible symptoms in the 7 days prior to survey participation. These symptoms included: fever or chills, cough, shortness of breath (or difficulty breathing), fatigue (or extreme tiredness), muscle or body aches or headache, congestion (or runny nose), sore throat, and loss of taste or smell, nausea or vomiting, and diarrhoea.

#### Non-pharmaceutical interventions (NPIs)

We extracted data on governments’ deployment of 8 containment measures from the Oxford Covid-19 Government Response Tracker (OxCGRT) project in each of the study countries^19^. These containment measures are:

- C1 Closing of schools and universities
- C2 Closing of workplace
- C3 Cancelling of public events
- C4 Limits on gatherings
- C5 Closing of public transport
- C6 Stay-at-home requirements
- C7 Restrictions on internal movement between cities/regions
- C8 Restrictions on international travel

With considerations of the potential implications of these rules on social contacts, and the changes in these rules over the study period in the 21 CoMix countries – e.g., government requirements on C4 and C5 did not vary in most study countries over the study period (see more in Supplementary Material I), we included C1, C2, C3, and C6 as potential factors of contacts at home, at work, and in other settings in our regression analyses. These restrictions were thoroughly reviewed by our international research team, and minor revisions were made where appropriate (details are provided on http://github.com/wongkerry/epipose_paper_1.git). We used a binary variable to denote if any recommendations or requirements were mandated in any parts of the country (0 = no measures were in place, and 1 = some recommendations or requirements were in place).

### Statistical analysis

R version 4.1.2 was used for all analyses, and the code and data are available online (see Data Availability Statement). The analyses conducted in this study are available on http://github.com/wongkerry/epipose_paper_1.git.

#### Descriptive

We calculated, for each country, summary statistics of the age, gender, household characteristics, risk perceptions towards catching and spreading COVID-19, use of face-covering, vaccination against COVID-19, self-reported high-risk status, and the presentation of symptoms of respiratory infection.

#### Mean number of contacts

We obtained the percentage of reported contacts in each setting – home, work, others, and in all settings – that were above 100 in each country; and calculated the crude mean number of contacts per person per day by censoring to 100 contacts. In previous studies, cut-off values for censoring of 100, 50 and 30 had been used since the mean number of contacts are heavily influenced by a few survey responses of very high number of contacts relative to the rest of the panel^15,16,20,21^. This analysis adopted the censoring value of 100 to capture the higher numbers of contacts provided (often in a work context at times of less stringent restrictions).

We calculated the mean and 95% confidence interval (CI) for the number of social contacts per participant per day over time in each study country, using bootstrapping with 1000 samples. Each participant was sampled with replacement and then all observations for selected participants were included in bootstrapped samples to account for dependency from repeated observation of the same participants. We calculated the mean number of contacts with a moving window over 2-week, overlapping intervals to increase the sample size per estimate and to include all participants from simultaneously running panels. Additionally, we assessed mean contacts in children aged <18 years. We stratified contacts in children by school attendance status (attended school versus did not attend school), and by age group in years – 0-4, 5-11, and 12-17 years.

#### Potential determinants of social contacts

We examined the effects that different potential determinants might have on social contacts in adult participants aged 18 years or above using an individual-level generalized additive mixed model (GAMM)^22^. The GAMM formulation also included a spline term of time, measured as year-month of the survey responses. For each of BE, DE, NL and UK, where data collection spanned the entire study period, we fitted separate setting-specific (home, work, other, and all settings) GAMM formulations, thus resulting in 16 separate models. In each model, we assumed reported contacts followed a zero-inflated Poisson distribution, with a random effect for participants and poststratification weights for age and gender based on the national population. The other 17 countries (in Groups 1, 2 and 3), where data collection took place over a shorter time period, were included in one single GAMM per setting, with a random effect for participants by country. A zero-inflated Poisson distribution and poststratification weights based on the national population were also applied. The poststratification weights were assigned by age groups 18-29, in 10-year age bands from 30 to 69, and 70 years old and above. We used the World Population Prospects 2019 standard projections overall and by gender for the 2020 national population in each country^23^. Participants with a missing age were not included in this analysis (0.3%). Weights for non-male and non-female participants were assigned by their age only. We additionally adjusted for the effect of time (measured as year-month), respondent fatigue (measured as the n-th survey response, per participant), and whether the survey response was provided for a weekday or at weekend.

## Results

The CoMix study collected data in 21 European countries between March 2020 and March 2022 (Figure 1). Except for the beginning and the end of the study, at least 10,000 survey responses were recorded across all participating countries on a monthly basis. At the highest point in January 2021, the number of survey responses recorded reached 37,808 in 14 countries.

**Figure 1.**
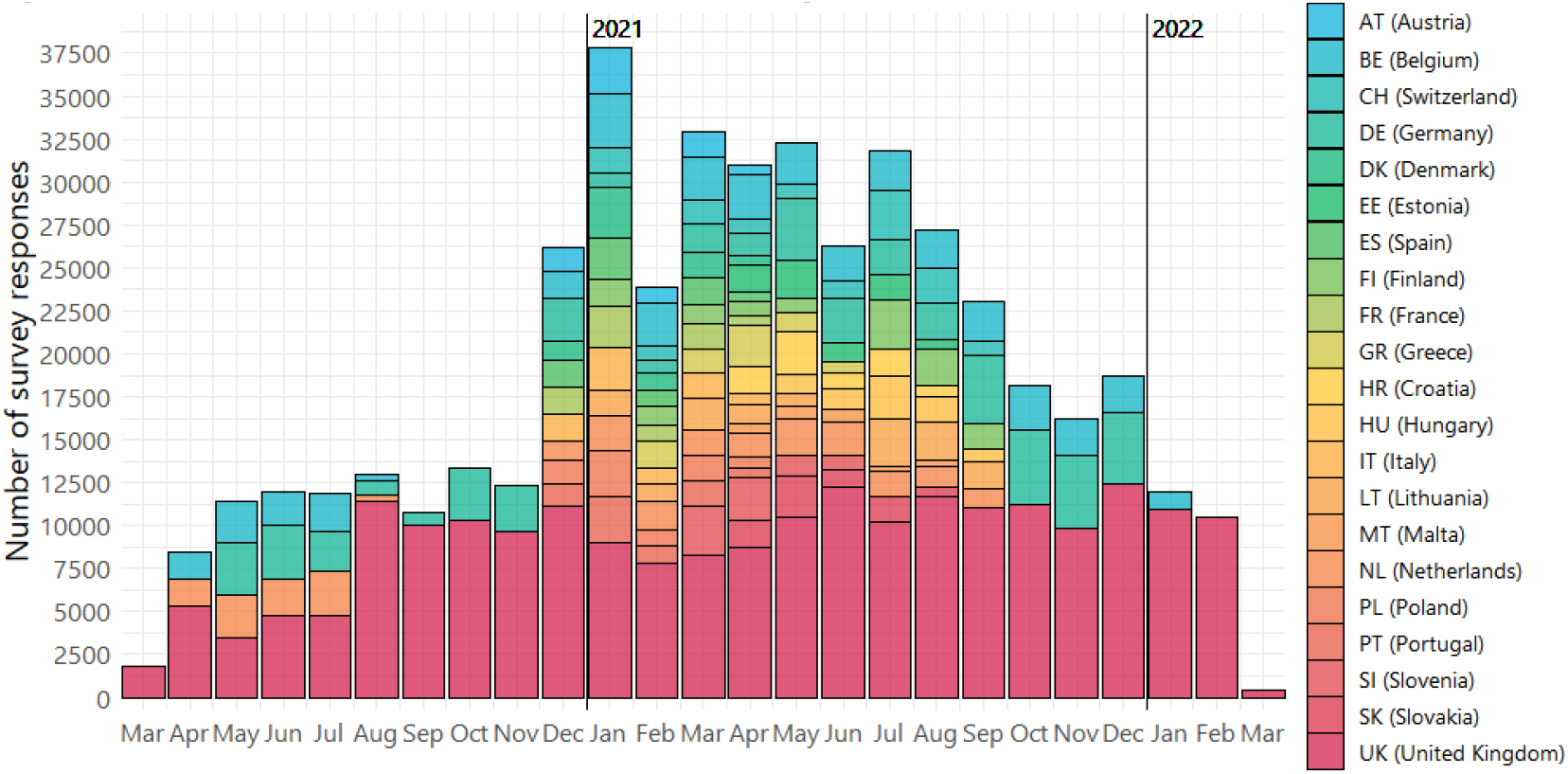
Timeline of data collection in participants aged 18 or above in 21 CoMix countries in Europe

Data was collected from 96,456 adults, who made a total of 463,336 responses to the CoMix survey (Table 1). Of these, 47% (217500) took place in the UK. Approximately 20% of the survey responses were given by participants aged between 40 and 59 years.

**Table 1.** Number of respondents, number of survey responses, and characteristics of survey responses aggregated over all survey responses

|  | All | UK | BE | NL | DE | G123<sup>+</sup> |
| --- | --- | --- | --- | --- | --- | --- |
| **No. respondents** | 96456 | 46779 | 4423 | 3643 | 6427 | 35184 |
| **No. responses** | 463336 | 217500 | 39620 | 24601 | 50018 | 131597 |
| **Age (%)** |  |  |  |  |  |  |
| 18-29 | 13.8 | 13.3 | 12.6 | 16.6 | 10.9 | 15.5 |
| 30-39 | 15.7 | 15.9 | 12.8 | 15.2 | 13.4 | 17.2 |
| 40-49 | 16.7 | 16.6 | 17.3 | 15.8 | 11.6 | 19.0 |
| 50-59 | 19.7 | 19.9 | 20.3 | 18.1 | 21.9 | 18.6 |
| 60-69 | 22.2 | 21.6 | 24.9 | 22.8 | 29.1 | 19.7 |
| 70 or above | 11.8 | 12.6 | 12.2 | 11.5 | 13.1 | 10.0 |
| **Gender (%)** |  |  |  |  |  |  |
| Female | 52.4 | 55.9 | 50.0 | 50.1 | 47.4 | 49.8 |
| Male | 47.6 | 44.1 | 50.0 | 49.9 | 52.6 | 50.2 |
| **Household characteristics** |  |  |  |  |  |  |
| Living with at least one 65+ years (%) | 19.3 | 18.8 | 20.9 | 17.8 | 20.5 | 19.6 |
| Mean household size | 2.3 | 2.3 | 2.2 | 2.1 | 2.1 | 2.6 |
| **Risk perception\* (%)** |  |  |  |  |  |  |
| Catching coronavirus | 5.3 | 4.2 | 4.2 | 5.3 | 4.7 | 7.7 |
| Serious illness from coronavirus | 16.4 | 14.3 | 19.9 | 25.2 | 19.8 | 16.1 |
| Spreading coronavirus to vulnerable persons | 22.0 | 19.4 | 20.2 | 26.3 | 26.8 | 24.2 |
| **Risk or risk mitigation (%)** |  |  |  |  |  |  |
| Used face covering | 64.0 | 58.2 | 61.4 | 34.3 | 75.8 | 75.4 |
| Vaccinated | 20.7 | 24.7 | 15.7 | 23.6 | 17.0 | 16.3 |
| Self-reported high-risk status | 30.3 | 30.1 | 27.1 | 36.0 | 38.1 | 27.7 |
| **Presentation of symptoms (%)** |  |  |  |  |  |  |
| Fever | 1.6 | 1.7 | 1.4 | 1.4 | 1.0 | 1.6 |
| Cough | 4.9 | 5.6 | 4.6 | 3.5 | 1.5 | 5.4 |
| Shortness of breath | 3.0 | 3.2 | 2.8 | 3.3 | 4.1 | 2.3 |
| Headache or body ache | 14.9 | 15.6 | 12.7 | 10.9 | 14.6 | 15.2 |
| Congestion | 8.0 | 8.0 | 7.9 | 7.7 | 7.1 | 8.3 |
| Sore throat | 4.4 | 5.1 | 4.1 | 3.7 | 3.3 | 3.9 |
| Fatigue or tiredness | 6.1 | 7.3 | 5.1 | 5.6 | 5.2 | 5.0 |
| Any symptoms<sup>++</sup> | 25.6 | 26.7 | 23.3 | 21.4 | 24.0 | 26.0 |

\* Participants answered a series of questions about their risk perception with Likert scale response options. Answers of "Strongly agree" are shown here.
<sup>+</sup> individual countries shown in Supplementary Material II
<sup>++</sup> any symptoms including fever, cough, shortness of breath, headache or body ache, congestion, sore throat, fatigue or tiredness, loss of taste or smell, and diarrhoea

Altogether, 19.3% of responses were provided by participants living with at least one other person aged 65 years or above. This proportion ranged between 14.5% in Switzerland and 33.2% in Italy (Supplementary Material II). Mean household size ranged between 2.0 people in Finland to 3.0 people in Poland (Supplementary Material II). Risk perception also differed between countries. The percentage reporting “strongly agree” to “I am likely to catch coronavirus” averaged 5.3%, and ranged between 2.1 % in Finland and 23.5% in Lithuania. About 10-25% were concerned about getting serious symptoms from coronavirus; and about 10-55% were concerned about spreading coronavirus to someone vulnerable.

Across all survey responses, self-reported usage of face-covering was 64.0% - lowest in the Netherlands (34.3%) and almost universal at 95% in Malta. Between <5% and 60% of survey responses were reported by partially or fully vaccinated participants. Approximately 20-40% of survey responses were reported by participants who self-identified as having at least one high-risk condition. Presentation of symptoms was reported in about one quarter of survey responses. The most common symptom was headache or body ache (14.9%).

### Crude mean number of contacts

Over 99.5% of daily contacts reported by adult participants were below 100. With censoring at 100, crude daily mean number of contacts were 3.22 (95%CI=3.19-3.26) in the UK over the entire study period (Table 2). In Belgium, Netherlands, and Germany, crude daily mean number of contacts after censoring were 3.94 (95%CI=3.85-4.03), 3.63 (95%CI=3.52-3.74), and 2.64 (95%CI=2.58-2.70), respectively. Among the 17 G123 countries, crude daily mean number of contacts varied considerably, and was highest in Malta (7.25), and lowest in Austria (2.81). Crude daily mean number of contacts without censoring are presented in Supplementary Material II.

**Table 2.** Crude daily mean number of contacts with censoring at 100 by setting and country

|  | UK | BE | NL | DE |  |  |  |
| --- | --- | --- | --- | --- | --- | --- | --- |
| **All** | 3.22 (3.19,3.26) | 3.94 (3.85,4.03) | 3.63 (3.52,3.74) | 2.64 (2.58,2.70) |  |  |  |
| **Home** | 1.16 (1.12,1.19) | 1.51 (1.42,1.60) | 1.20 (1.09,1.31) | 0.96 (0.91,1.02) |  |  |  |
| **Work** | 1.07 (1.03,1.10) | 1.45 (1.36,1.54) | 1.46 (1.35,1.57) | 0.62 (0.56,0.68) |  |  |  |
| **Others** | 1.11 (1.07,1.14) | 1.12 (1.03,1.21) | 1.08 (0.97,1.19) | 0.74 (0.68,0.80) |  |  |  |
|  | ES | FR | IT | AT | DK | PO | PL |
| **All** | 3.09 (2.99,3.19) | 3.15 (2.98,3.32) | 3.03 (2.89,3.17) | 2.81 (2.65,2.97) | 3.72 (3.55,3.89) | 3.97 (3.79,4.16) | 3.94 (3.75,4.13) |
| **Home** | 1.69 (1.59,1.80) | 1.23 (1.07,1.40) | 1.60 (1.47,1.74) | 1.33 (1.17,1.49) | 1.30 (1.13,1.48) | 1.76 (1.58,1.95) | 1.68 (1.49,1.87) |
| **Work** | 0.90 (0.80,1.01) | 1.06 (0.89,1.22) | 0.91 (0.77,1.04) | 0.82 (0.66,0.98) | 1.17 (1.00,1.35) | 1.15 (0.96,1.33) | 1.08 (0.90,1.27) |
| **Others** | 0.53 (0.43,0.63) | 0.92 (0.75,1.09) | 0.58 (0.44,0.71) | 0.71 (0.55,0.87) | 1.28 (1.11,1.45) | 1.24 (1.06,1.42) | 1.26 (1.07,1.45) |
|  | CH | FI | LT | GR | SL |  |  |
| **All** | 4.16 (3.98,4.33) | 3.84 (3.65,4.02) | 4.69 (4.50,4.88) | 3.50 (3.34,3.67) | 4.31 (4.09,4.53) |  |  |
| **Home** | 1.31 (1.13,1.48) | 1.01 (0.83,1.20) | 1.30 (1.12,1.49) | 1.63 (1.47,1.80) | 1.87 (1.65,2.09) |  |  |
| **Work** | 1.72 (1.55,1.89) | 1.66 (1.47,1.84) | 1.67 (1.48,1.85) | 1.05 (0.88,1.21) | 1.36 (1.14,1.58) |  |  |
| **Others** | 1.34 (1.16,1.51) | 1.40 (1.21,1.58) | 1.99 (1.80,2.18) | 0.93 (0.77,1.09) | 1.32 (1.10,1.54) |  |  |
|  | EE | HU | SK | MT | HR |  |  |
| **All** | 5.25 (4.98,5.52) | 5.74 (5.49,6.00) | 4.55 (4.30,4.80) | 7.25 (6.78,7.71) | 3.75 (3.56,3.94) |  |  |
| **Home** | 1.35 (1.08,1.61) | 1.93 (1.68,2.18) | 1.62 (1.37,1.87) | 2.19 (1.72,2.65) | 1.58 (1.39,1.77) |  |  |
| **Work** | 2.14 (1.89,2.39) | 2.09 (1.82,2.35) | 1.65 (1.40,1.90) | 2.26 (1.79,2.72) | 1.28 (1.09,1.47) |  |  |
| **Others** | 2.07 (1.80,2.34) | 1.87 (1.62,2.12) | 1.59 (1.34,1.84) | 3.10 (2.64,3.57) | 1.06 (0.87,1.25) |  |  |

#### Social contacts over time

Across the study period in all countries, the lowest number of daily mean number of contacts of approximately 2 was recorded in Germany for most parts of the study period (Figure 2), and the highest contact of 9 was recorded in Malta in April 2021. In UK, Belgium, Germany, Netherlands, Italy, Poland, and Finland, where data were available, mean daily contacts during the pandemic dropped far below the pre-pandemic level as measured in the POLYMOD study^11^. In the most extreme case, in Italy, mean daily contacts were between 2.2 (95%CI=2.0-2.5) to 3.7 (95%CI=3.4-4.1), as opposed to 19.8 in 2005-06^11^. Changes over time were primarily driven by a large reduction in contacts at work and in other settings, as home contacts remained broadly static in all countries. In the UK, Belgium and the Netherlands where data were available, an increase in contacts at work was somewhat gradual after the first several months of the pandemic in 2020, as opposed to a more rapid upsurge for contacts in other settings (Figure 2).

**Figure 2.**
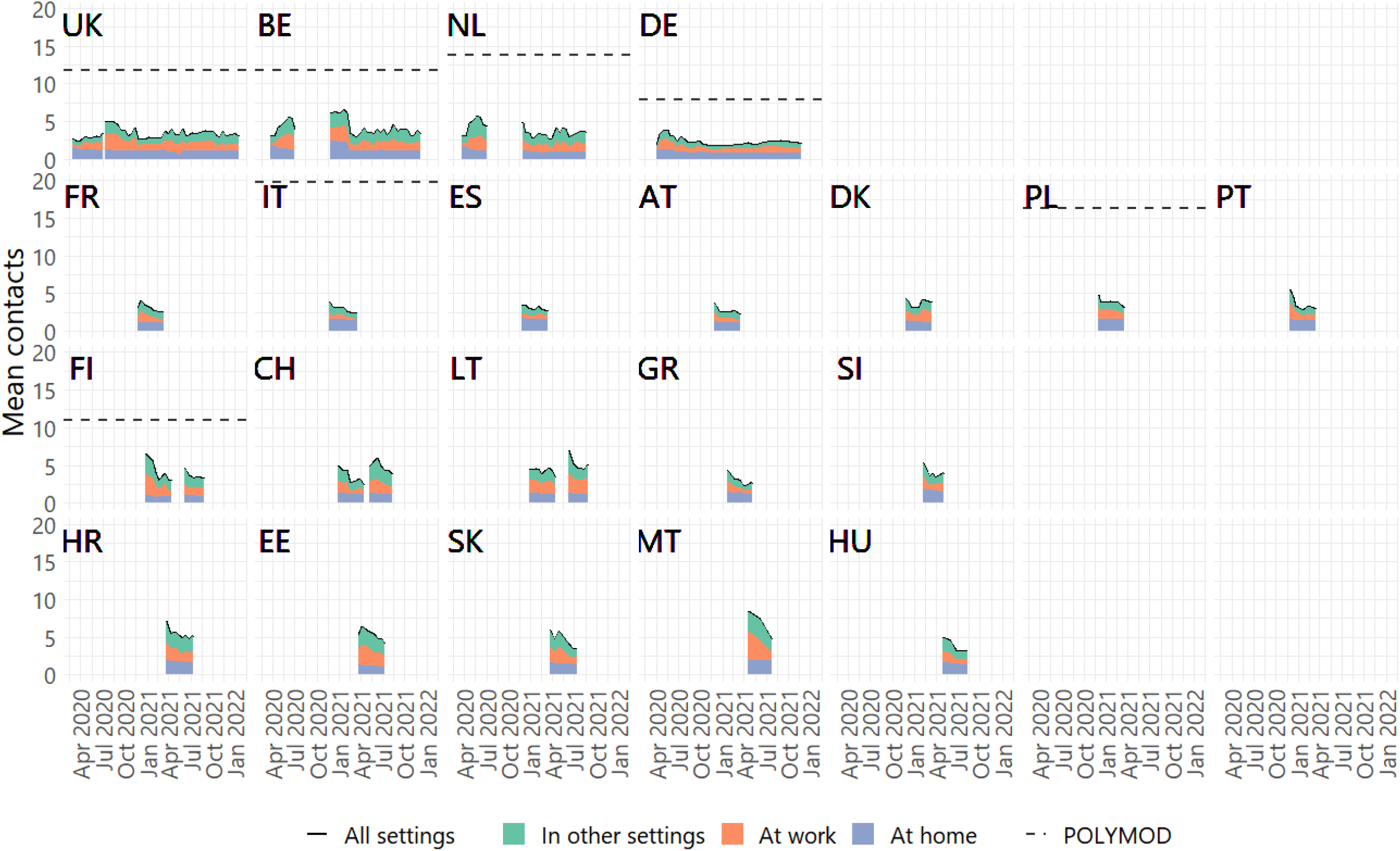
Mean number of contacts by settings over time, with mean from POLYMOD^11^ where data is available

### Factors associated with social contacts at home, at work, and in other settings

We compared the relative difference in mean contacts brought about by difference factors using GAMM models (Figure 3). Compare to the reference age group of 18-29 years, those aged 70 or above reported 0.61 (95%CI=0.59-0.62) to 0.84 (95%CI=0.82-0.86) times the contacts in all settings across countries. Compared to women, men consistently reported fewer contacts at home in all countries, and fewer contacts at work in all countries apart from the Netherlands (Suppl IV).

**Figure 3.**
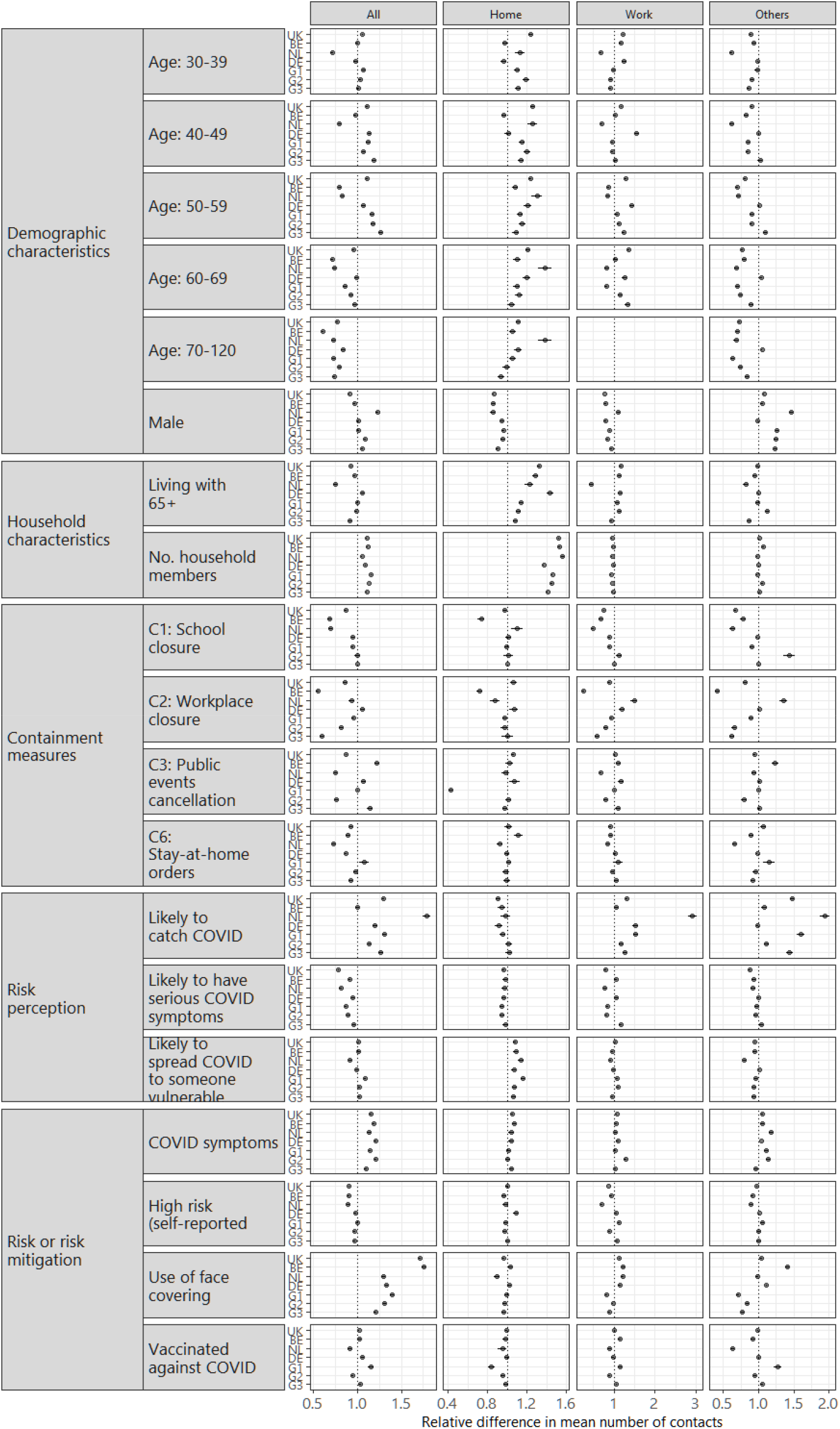
Forest plot showing the relative differences on mean number of contacts per person in UK, NL, DE, BE, and 17 Groups 1, 2 and 3 countries, by settings (home, work, other, and all settings). Relative differences pooled across the seven locations are also included.

Household size was associated with more contacts at home, but had little to no effect on contacts at work and in other settings. The effect of living with at least one person aged 65+ varied across countries (Figure 3).

The associations between social contacts and most of the included government restrictions differed across countries. Cancellation of public events, for instance, was associated with making 0.43 times (95%CI=0.41-0.45) fewer contacts at home in Group 1 countries (where data collection took place between December 2020 and April 2021), and had minimal effect in other locations. Workplace closure was not only associated with fewer contacts at work, but also in other settings in UK, and Group 2 and Group 3 countries. Contacts in settings other than home and work appeared lower under stay-at-home orders in all locations other than in the UK and in Group 1 countries.

Participants who strongly agreed that they may get COVID-19 reported more contacts in all settings (ranged from 1.14 (95%CI=1.12-1.16) times in Group 2 countries to 1.79 (95%CI=1.75-1.84) times in the Netherlands), other than in Belgium (1.00, 95%CI=0.98-1.03). People who strongly agreed that they might get severely sick from COVID-19 reported fewer contacts – the relative differences ranged from 0.79 (95%CI=0.78-0.79) to 0.96 (95%CI=0.94-0.98). Concerns about spreading COVID-19 to someone vulnerable was associated with more contacts at home.

Participants who reported COVID-19 symptoms tended to report more contacts at work (of 1.02 (95%CI=1.00-1.04) to 1.28 (95%CI=1.25-1.31)). Except for Belgium, COVID-19 symptoms were also associate with more contacts in other settings (1.04 (95CI=1.04-1.05) to 1.18 (95%CI=1.15-1.21)). High risk status and vaccination status appeared to have minimal effect. The use of face covering was associated with 1.20 (95%CI=1.19-1.22) to 1.75 (95%CI=1.73-1.77) times more contacts in all settings.

The number of contacts reported appeared to be negatively associated with the number of surveys responded to, per participant (Figure 4). The relative difference in the mean number of contacts across all settings declined most dramatically for Groups 1, 2, and 3 countries, where the survey sample was not replenished. By the fifth survey round, for instance, participants reported approximately 60% of the number of contacts reported at their first round of survey participation.

**Figure 4.**
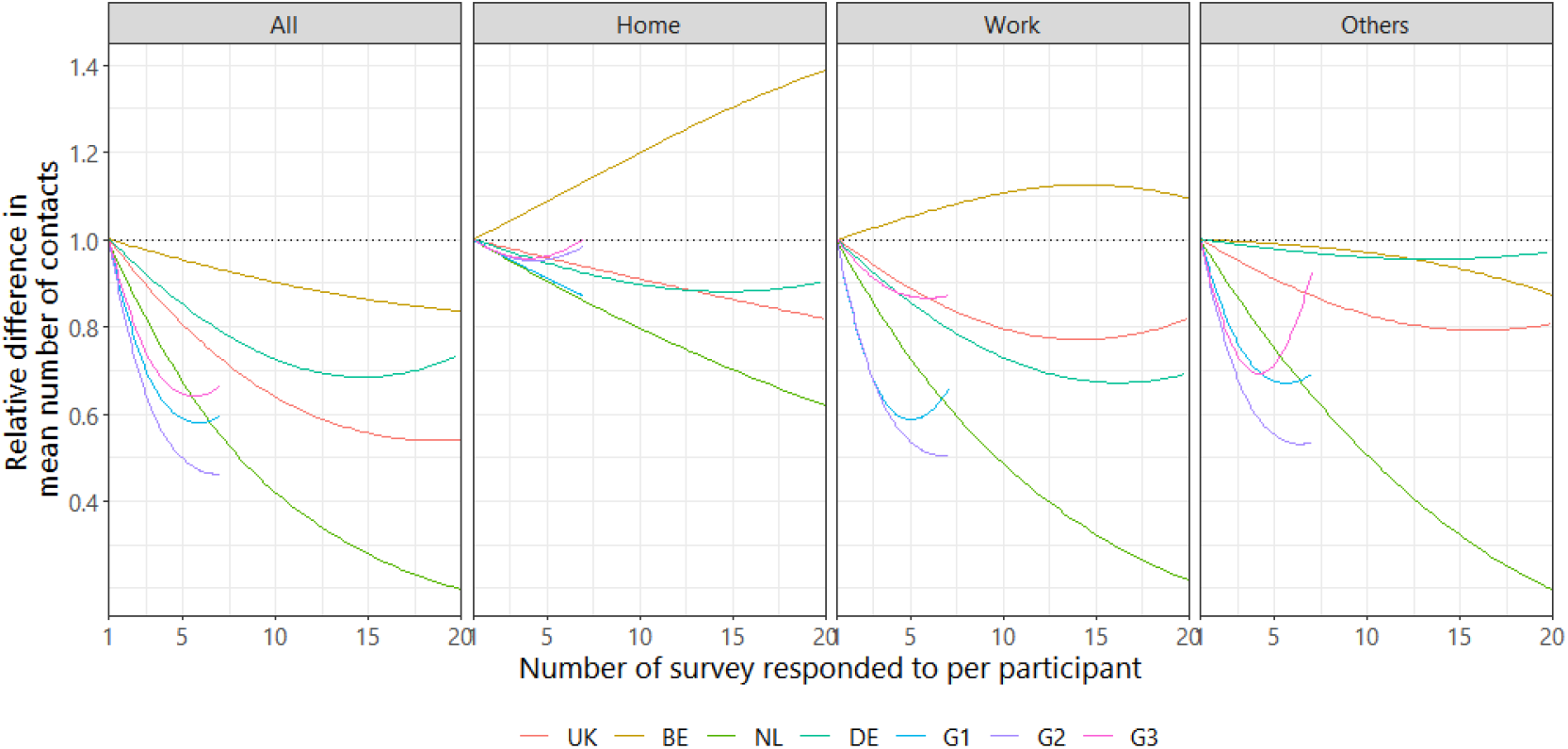
Respondent fatigue effect captured as the number of surveys responded to per participant.

### Contacts in children by school attendance status

Among children who attended school, the mean daily reported number of contacts across all countries ranged between approximately 5 to 15 (Figure 5). By age, mean daily contacts in children aged 0-4 years was 8.1 across countries, and was the most similar to POLYMOD (10.2). Mean reported contacts were 10.4 for children aged 5-11 years and 12-17 years, respectively, and was somewhat lower than POLYMOD (14.8 and 17.6). Children who did not go to schools made far fewer contacts (average of less than 5 in most countries).

**Figure 5.**
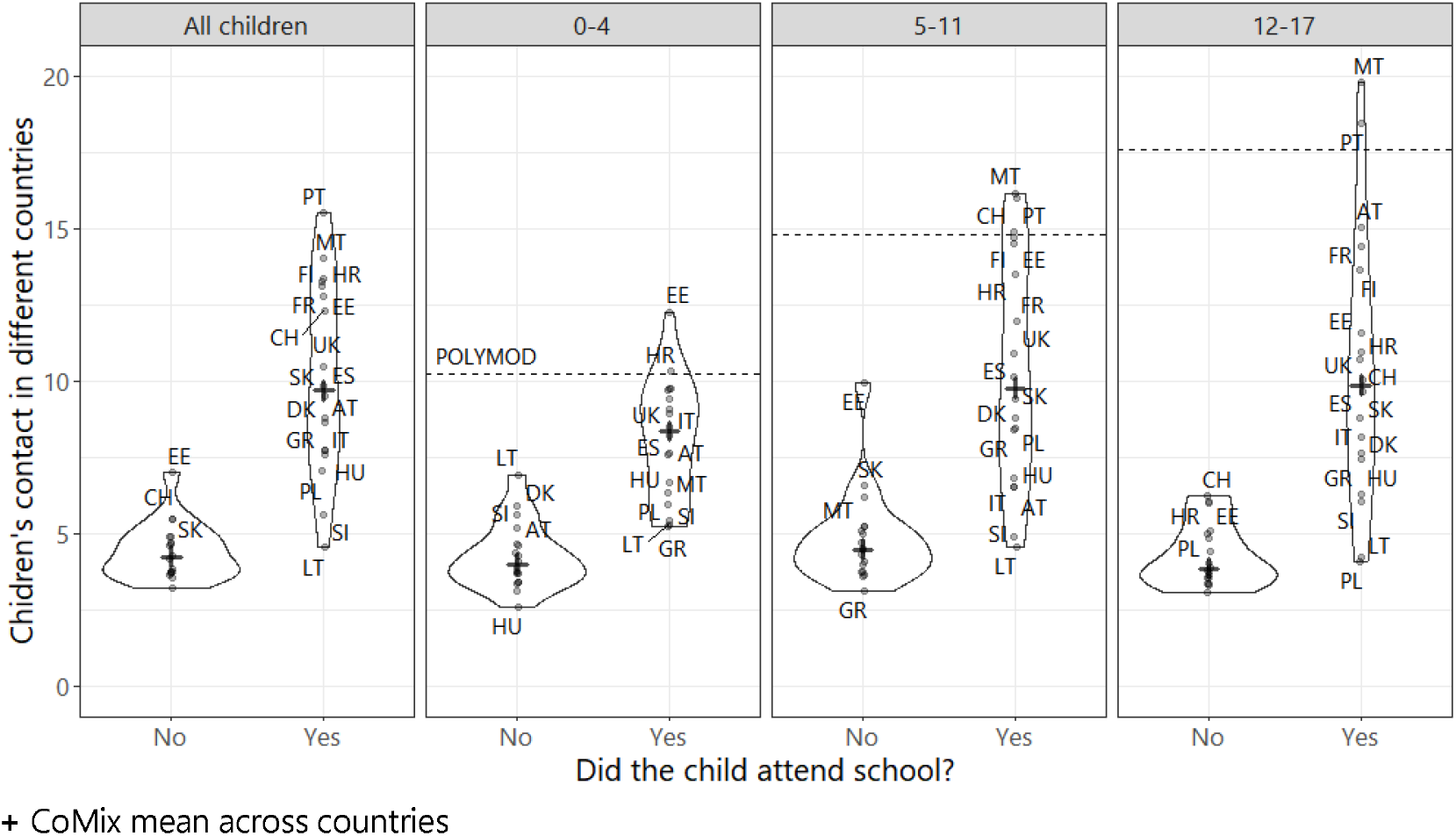
Contacts in children aged <18 in all study countries by school attendance status

## Discussion

To our knowledge, this study on social contact during the pandemic has been the largest of its kind in terms of the number of countries, participants, and observations included. The CoMix survey, collecting standardized repeated cross-sectional data, facilitated an examination of social contacts across 21 European countries over a 24-month period, and assessment contact rates before and during COVID-19 using established findings, where possible. More than 460,000 survey responses were considered in the current analysis. In all examined countries in the region, despite changes in NPIs, mean contact rates remained low over the study period. In the subset of countries where data were available, mean contact rates were much lower than pre-pandemic levels, predominately due to fewer contacts not at home. NPIs showed clear immediate effects far beyond the time when many different restrictions were lifted. The relationships between individual perceptions or national policy determining contacts varied across the countries studied.

Easing, or partial easing, of the first round of major NPIs began in May 2020 in the countries represented in this study. Much of the increase in social contacts around that time was largely based on more contacts at work^16^. Findings from our study across different countries confirmed that containment measures at workplaces and stay-at-home orders were both associated with fewer contacts in the population. Following the lifting of restrictions, the initial increases in work contacts were relatively slow, and possibly influenced by sector of employment, availability of face covering, and other adjustments enabling the return to the workplace. The pandemic has resulted in changes to the working arrangements of many who are now based at home and may continue to work at home, in some capacity, for the foreseeable future. In this study, we also found lower contacts in non-work settings under workplace closure mandates, suggesting potential wider effects on social contacts outside of work. Conversations at work with colleagues may offer opportunities for social connections beyond work, and the shift to homeworking potentially reduces the number of opportunities for these spontaneous interactions^24^. The full consequences of working at home on social contacts, especially the unintended, knock-on effect in non-work related social contacts (e.g. increased risk of feeling lonely^25^), are yet to be carefully examined.

Our study also identified a link between contacts at home and concerns about spreading COVID-19 to vulnerable persons. Such perception was not observed for contacts outside the home. Whilst measures focused on social distancing behaviours can be undertaken to reduce the spread of COVID-19 within the community, people with symptoms of the virus were instructed or advised to remain in their homes. Within-household contacts are thought to be the dominant route of disease spread when extensive community control measures are in place^26^. Despite guidance advising household members to socially distance, individuals are likely to interact repeatedly. Those living with household members who are older, have underlying medical conditions, or share a bed might be particularly concerned^27–29^. Strategies to support people quarantining at home with other household members remain vital in the ongoing pandemic.

Whilst we set out to identify determinants of social contacts, our study – observational in nature – might have picked up on other correlates of social contacts. In most countries, presentation of symptoms, use of face-covering, vaccination against COVID-19, and concerns about infection (but not getting severely ill) showed associations with more contacts in non-home settings. People who had more contacts likely had a greater tendency to acquire COVID-19, were concerned about it, and might have adopted the behaviour of using face-covering – or had to wear one to go to work, and to travel on public transport. Our finding on greater use of face-covering aligns with previous studies in the US^30^ and Japan^31^ showing masks-wearing was more prominent among individuals who were more socially integrated and who spent less time at home, including going to commercial locations and restaurants.

The current study offers important insights into the extent to which children meet others when they do not attend school. Whilst contact rates were lower when children did not go to school, they were still, on average, making contacts with more people than adult. Concerns as to grandparents or older people taking care of children when they are not at school have been raised^32^. Given that school closures are often accompanied by advice to parents to limit the contact their children have with others, further research into the characteristics of children’s contacts on days they do not go to school may be helpful in improving the advice that is given out.

Our findings should be interpreted with several limitations in mind. First, there were likely differences in the extent to which the NPIs were implemented in different countries. For instance, the allowable reasons to leave one’s home during periods of stay-at-home orders might vary across countries. Our measure on NPIs did not capture these variabilities, as well as any subnational differences. Second, CoMix data collection had different temporal coverages across the study countries, including periods of higher social activities such as end of year holiday period. We conducted analyses grouping countries with field work at similar times to partially account for the effect of time. Third, our grouping of countries based on field work time periods might have put countries with different characteristics and measures together, thus potentially masking country-specific relationships. For instance, perception and acceptability towards mask usage across countries or cultures might vary, leading to different relationships with contact rates. Fourth, survey fatigue appeared substantial in our study. As contact surveys continue to be used as an important public health tool for emerging infectious disease, survey methodologies may need to be adjusted. Limiting the maximum number of survey response per participant, and using fewer, but strategically timed survey rounds may yield results that more accurately capture contact patterns in the population. In addition, efforts to minimize response bias is likey more important now than during pre-pandemic times – such as when POLYMOD was conducted – since the relationship between contacts and disease transmission has become more widely acknowledged in the population. Lastly, the effect of some covariates may be better uncovered with longitudinal models. For instance, individuals may make more contacts when they are vaccinated compared to before, but this overall effect could be masked by other covariates when the individual level of the data is not accounted for. Such changes are beyond the scope of the current paper, and might be better addressed at the level of individual person, with approaches such as paired measures^33^.

## Conclusion

The COVID-19 pandemic has led to an unprecedented global impact with wide ranging health and social consequences. Across 21 countries in the European region, we found that in-person contacts remained far below pre-pandemic levels throughout the two-year study period. In this multi-country analysis, we presented a detailed historical record of social behaviour during the COVID-19 pandemic, research insights on the influence of NPIs, the key roles of individuals’ perceived pandemic threat level, and personal circumstances, and insights the can be gained by comparing the differences between countries. Our study provides an account across different countries, and paves the way to develop future studies, coordinated at the regional level, to guide infectious diseases outbreak responses, particularly for a large-scale pandemic.

## Data Availability

http://github.com/wongkerry/epipose_paper_1.git

## Acknowledgement

The following authors were part of the CoMix Europe Working Group. Each contributed to consultation for policy input, interpretation of data and findings, and supported the drafting of the manuscript, and approved the work for publication:

- Daniela Paolotti, Michele Tizzani and Ciro Cattuto (Data Science for Social Impact, ISI Foundation)
- Andrea Schmidt, Gerald Gredinger, and Sophie Stumpfl (Health Economics and Health System Analysis, Austrian National Public Health Institute)
- Joaquin Baruch, and Tanya Melillo (Infectious Disease Prevention & Control Unit, Ministry for Health, Malta)
- Henrieta Hudeckova, Jana Zibolenova, and Zuzana Chladna (Department of Public Health, Comenius University Bratislava, Jessenius Faculty of Medicine in Martin)
- Magdalena Rosinska, and Marta Niedzwiedzka-Stadnik (Department of Infectious Disease Epidemiology and Surveillance, National Institute of Public Health NIH - National Research Institute, Poland)
- Krista Fischer, Sigrid Vorobjov, and Hanna Sõnajalg (Department of drug and infectious diseases epidemiology, National Institute for Health Development, Estonia)
- Christian Althaus, Nicola Low, and Martina Reichmuth (Institute of Social and Preventive Medicine, University of Bern)
- Kari Auranen, and Markku Nurhonen (Department of Health Security and Dept. of Public Health and Welfare, National Institute for Health and Welfare, Finland)
- Goranka Petrovic, and Zvjezdana Lovric Makaric (Division for Epidemiology of Communicable Diseases, Croatian Institute of Public Health)
- Sónia Namorado, Constantino Caetano, and Ana João Santos (Department of Epidemiology, National Institute of Health Dr Ricardo Jorge, Portugal)
- Gergely Röst, Beatrix Oroszi, and Márton Karsai (Bolyai Institute, University of Szeged)
- Mario Fafangel, Petra Klepac, and Natalija Kranjec (Communicable Diseases Centre, National Institute of Public Health, Slovenia)
- Cristina Vilaplana, and Jordi Casabona (Experimental Tuberculosis Unit, Institut d’Investigació en Ciències de la Salut Germans Trias i Pujol, Spain)

We acknowledge support from the European Centre for Disease Prevention and Control (ECDC) in setting up the collaborations between the Epipose consortium, and universities and public health institutions in all other countries. We gratefully acknowledge the tremendous efforts put in all the steps by the EpiPose consortium, its collaborators and Ipsos.

We would also like to thank the team at Ipsos who have been excellent in running the survey, collecting the data, and allowing for this study to, happen at a rapid speed. We acknowledge the exceptional project management support given by Sarah Vercruysse and Bieke Vanhoutte.

## Authors’ contributions

KLMW, CIJ and WJE conceived of and planned the analysis; KLMW performed the main analysis based on methods previously developed by AG, CIJ, and WJE; PC and NH provided comments and discussions on the analytical methods. CIJ and WJE designed the CoMix contact survey. Data management and curating was performed by PC (Belgium), VKJ and AK (Germany), and CIJ, AG, and KLMW (UK). All authors reviewed and approved the manuscript. The CoMix Europe Working Group provided discussion and comments.

## Funding

The following funding sources are acknowledged as providing funding for the named authors. HPRU in Modelling & Health Economics (NIHR200908: KLMW); European Union Horizon 2020 research and innovation programme (EpiPose 101003688: AG, WJE); European Research Council under the European Union Horizon 2020 research and innovation programme (TransMID 682540: CF, PB, NH) This research was partly funded by the Global Challenges Research Fund (GCRF) project RECAP managed through RCUK and ESRC (ES/P010873/1: CIJ) NIHR (PR_OD_1017_20002: WJE) UK MRC (MC_PC_19065 - Covid 19: Understanding the dynamics and drivers of the COVID-19 epidemic using real-time outbreak analytics: WJE).

In Belgium, CoMix data collection in Belgium was made possible with financial support of Janssen Pharmaceuticals and the national public health institute of Belgium, Sciensano.

In Germany, the COVIMOD project is funded by intramural funds of the Institute of Epidemiology and Social Medicine, University of Münster, and of the Institute of Medical Epidemiology, Biometry and Informatics, Martin Luther University Halle-Wittenberg, as well as by funds provided by the Robert Koch Institute, Berlin, the Helmholtz-Gemeinschaft Deutscher Forschungszentren e.V. via the HZEpiAdHoc “The Helmholtz Epidemiologic Response against the COVID-19 Pandemic” project, the Saxonian COVID-19 Research Consortium SaxoCOV (co-financed with tax funds on the basis of the budget passed by the Saxon state parliament), the Federal Ministry of Education and Research (BMBF) as part of the Network University Medicine (NUM) via the egePan Unimed project (funding code: 01KX2021) and the Deutsche Forschungsgemeinschaft (DFG, German Research Foundation, project number 458526380).

The funders had no role in study design, analysis, decision to publish or preparation of the manuscript.

**Supplementary Material I.**
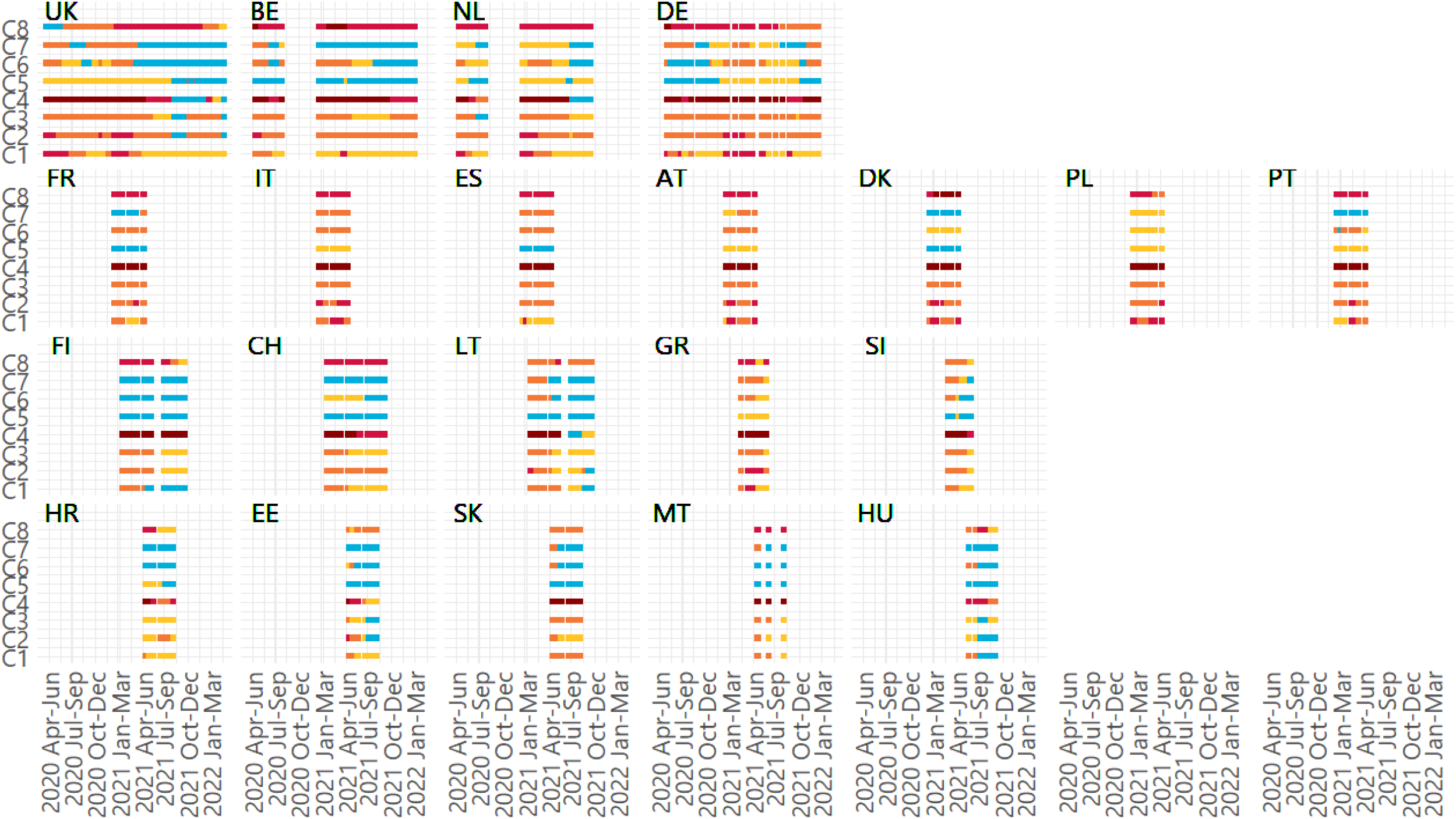
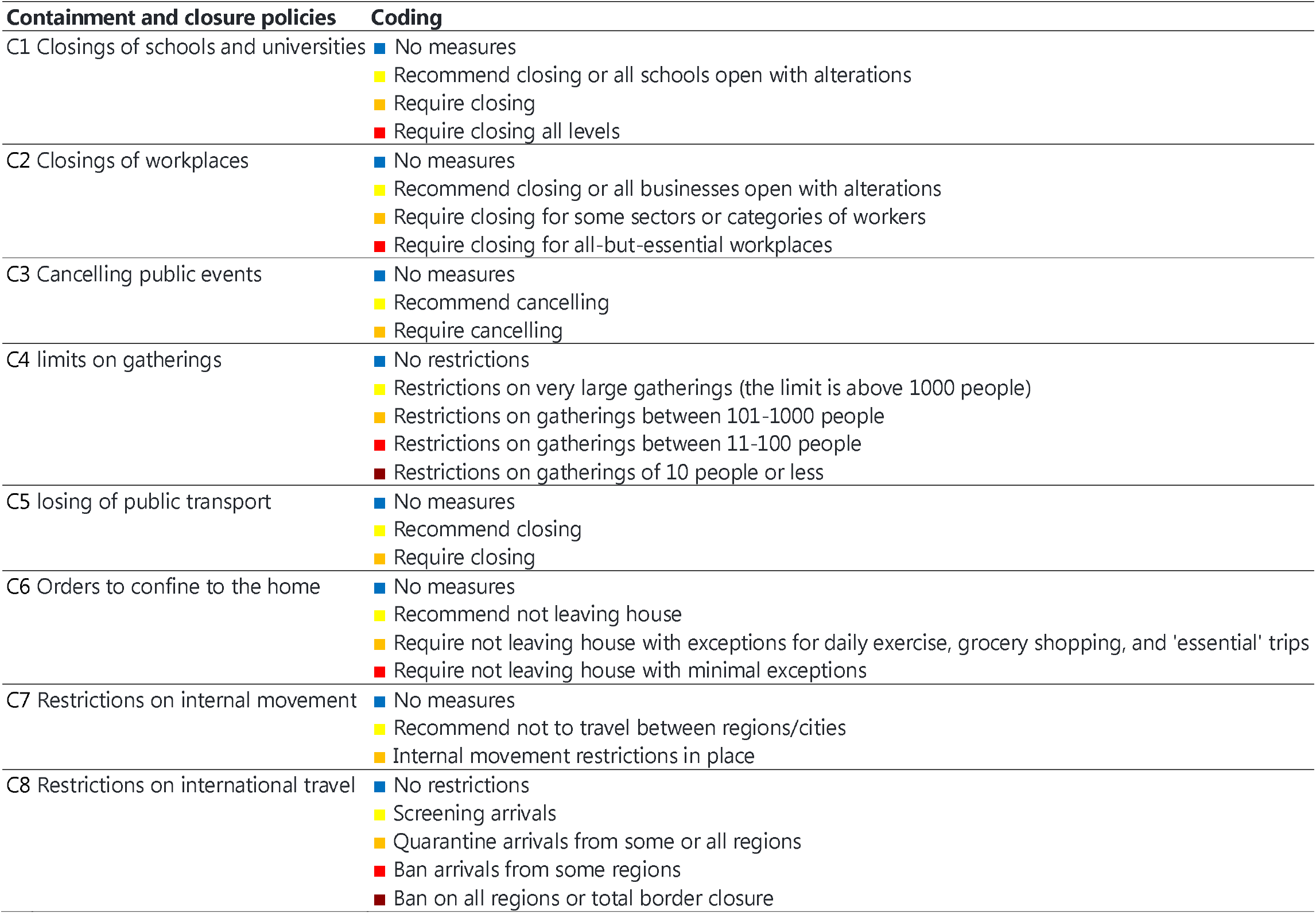

**Supplementary Material II.** Characteristics of survey responses in 17 G123 countries

|  | at | ch | dk | ee | es | fi | fr | gr | hr | hu | it | lt | mt | pl | pt | si | sk |
| --- | --- | --- | --- | --- | --- | --- | --- | --- | --- | --- | --- | --- | --- | --- | --- | --- | --- |
| respondents | 1737 | 4425 | 1728 | 1852 | 1622 | 2622 | 1704 | 2240 | 1971 | 2078 | 1616 | 2593 | 1702 | 1724 | 1839 | 1998 | 1733 |
| responses | 7025 | 12149 | 7044 | 6967 | 7048 | 11784 | 7025 | 7111 | 7080 | 7207 | 7048 | 12763 | 2759 | 7011 | 7018 | 7510 | 7048 |
| 18-29 | 18.5 | 15.6 | 14.1 | 13.1 | 14.6 | 14.8 | 14.9 | 16.1 | 18.4 | 12.4 | 15.0 | 14.9 | 17.3 | 18.8 | 16.0 | 15.8 | 15.3 |
| 30-39 | 17.2 | 18.5 | 10.5 | 16.9 | 16.9 | 15.3 | 17.4 | 21.1 | 16.4 | 16.9 | 13.5 | 16.5 | 24.8 | 19.3 | 16.1 | 20.3 | 20.5 |
| 40-49 | 16.7 | 16.3 | 13.7 | 16.6 | 22.5 | 15.5 | 16.5 | 26.3 | 20.1 | 22.2 | 21.8 | 16.9 | 20.8 | 19.2 | 21.8 | 21.4 | 20.6 |
| 50-59 | 19.6 | 20.2 | 22.5 | 18.4 | 17.7 | 17.1 | 15.3 | 24.4 | 18.1 | 16.5 | 15.7 | 18.7 | 16.2 | 18.0 | 19.2 | 20.9 | 16.5 |
| 60-69 | 18.5 | 17.2 | 22.8 | 18.7 | 18.8 | 23.0 | 22.4 | 10.1 | 21.6 | 22.4 | 20.5 | 23.4 | 14.5 | 18.9 | 18.4 | 18.1 | 19.9 |
| 70-120 | 9.5 | 12.1 | 16.4 | 16.3 | 9.4 | 14.3 | 13.4 | 2.0 | 5.5 | 9.7 | 13.5 | 9.6 | 6.5 | 5.8 | 8.5 | 3.5 | 7.2 |
| female | 47.0 | 49.0 | 47.2 | 54.2 | 49.6 | 47.2 | 46.6 | 48.7 | 51.4 | 47.4 | 46.6 | 52.9 | 57.8 | 55.6 | 45.6 | 56.8 | 47.4 |
| male | 53.0 | 51.0 | 52.8 | 45.8 | 50.4 | 52.8 | 53.4 | 51.3 | 48.6 | 52.6 | 53.4 | 47.1 | 42.2 | 44.4 | 54.4 | 43.2 | 52.6 |
| old | 14.7 | 14.5 | 17.4 | 19.8 | 25.7 | 16.8 | 19.8 | 15.1 | 23.6 | 20.7 | 33.2 | 18.9 | 17.4 | 20.8 | 21.2 | 17.0 | 20.8 |
| hh_size | 2.3 | 2.3 | 2.1 | 2.3 | 2.8 | 2.0 | 2.4 | 2.8 | 2.9 | 2.6 | 2.8 | 2.5 | 3.0 | 3.0 | 2.6 | 2.8 | 2.7 |
| likely | 3.1 | 4.9 | 2.8 | 9.8 | 6.0 | 2.1 | 10.4 | 15.7 | 7.2 | 4.1 | 3.6 | 23.5 | 6.0 | 6.9 | 9.5 | 4.5 | 2.8 |
| serious | 10.5 | 9.3 | 12.8 | 20.0 | 26.1 | 16.8 | 14.3 | 18.1 | 14.6 | 13.3 | 15.6 | 17.7 | 19.6 | 22.5 | 23.9 | 13.9 | 9.4 |
| spread | 11.2 | 14.8 | 29.6 | 13.7 | 55.4 | 13.3 | 25.0 | 30.4 | 30.2 | 11.1 | 24.2 | 25.9 | 30.2 | 23.2 | 50.3 | 23.4 | 16.7 |
| mask | 68.2 | 76.7 | 74.4 | 49.9 | 90.5 | 65.3 | 84.9 | 89.8 | 85.3 | 36.8 | 89.0 | 75.3 | 94.6 | 85.5 | 83.8 | 72.5 | 78.4 |
| vacc | 3.8 | 20.3 | 3.5 | 30.7 | 2.8 | 25.3 | 5.6 | 11.2 | 22.4 | 35.1 | 3.7 | 24.1 | 60.5 | 4.2 | 2.5 | 10.7 | 23.3 |
| risk | 22.2 | 22.7 | 22.9 | 32.5 | 31.8 | 31.7 | 25.9 | 19.2 | 28.5 | 33.1 | 21.7 | 33.8 | 19.7 | 35.6 | 26.0 | 24.7 | 31.0 |
| symp_fever | 1.0 | 1.6 | 1.2 | 1.4 | 1.6 | 2.2 | 2.3 | 1.5 | 1.9 | 1.6 | 1.7 | 1.5 | 0.8 | 2.6 | 0.8 | 1.6 | 1.5 |
| symp_cough | 5.4 | 4.9 | 7.0 | 3.7 | 7.8 | 5.2 | 5.1 | 4.6 | 4.9 | 6.6 | 4.9 | 3.6 | 1.6 | 10.1 | 5.2 | 6.1 | 4.5 |
| symp_sob | 2.3 | 2.6 | 3.0 | 1.8 | 2.5 | 3.0 | 2.7 | 1.6 | 2.2 | 2.3 | 2.2 | 1.6 | 1.2 | 2.9 | 1.6 | 2.3 | 3.4 |
| symp_ache | 14.8 | 13.8 | 16.9 | 13.9 | 21.9 | 16.8 | 15.8 | 13.7 | 12.9 | 16.7 | 15.6 | 10.2 | 12.7 | 23.0 | 14.6 | 12.2 | 14.7 |
| symp_congestion | 10.5 | 6.5 | 9.5 | 8.3 | 9.2 | 14.2 | 7.5 | 6.1 | 7.3 | 7.5 | 6.0 | 6.3 | 4.0 | 13.5 | 7.4 | 8.6 | 5.2 |
| symp_sore_throat | 3.8 | 3.5 | 4.2 | 2.9 | 5.4 | 3.7 | 4.5 | 3.1 | 2.9 | 3.8 | 4.8 | 3.2 | 1.6 | 6.9 | 3.4 | 3.8 | 4.1 |
| symp_tired | 6.4 | 5.3 | 4.5 | 4.2 | 4.5 | 5.3 | 4.4 | 5.3 | 3.7 | 6.7 | 4.2 | 4.3 | 3.6 | 8.5 | 3.3 | 4.0 | 5.7 |
| symp_any | 28.6 | 24.8 | 28.4 | 24.6 | 34.4 | 30.1 | 27.1 | 23.6 | 22.6 | 27.2 | 26.6 | 18.3 | 18.9 | 37.6 | 23.5 | 22.4 | 23.8 |

**Supplementary Material III.** Crude daily mean daily number of contacts without censoring by setting and country

|  | UK | BE | NL | DE |  |  |  |
| --- | --- | --- | --- | --- | --- | --- | --- |
| **All** | 4.01 (3.86,4.16) | 5.36 (4.75,5.97) | 5.39 (3.75,7.03) | 2.95 (2.77,3.12) |  |  |  |
| **Home** | 1.16 (1.01,1.30) | 1.51 (0.90,2.13) | 1.20 (-0.44,2.84) | 0.96 (0.79,1.14) |  |  |  |
| **Work** | 1.31 (1.16,1.46) | 2.01 (1.40,2.63) | 2.38 (0.74,4.02) | 0.62 (0.45,0.80) |  |  |  |
| **Others** | 1.55 (1.40,1.70) | 1.84 (1.22,2.45) | 1.81 (0.17,3.45) | 0.81 (0.63,0.98) |  |  |  |
|  | ES | FR | IT | AT | DK | PO | PL |
| **All** | 3.20 (3.01,3.40) | 3.41 (3.03,3.80) | 3.13 (2.94,3.32) | 2.99 (2.74,3.24) | 3.93 (3.60,4.27) | 4.98 (3.87,6.09) | 5.19 (3.83,6.54) |
| **Home** | 1.69 (1.50,1.88) | 1.23 (0.85,1.62) | 1.60 (1.41,1.80) | 1.33 (1.08,1.58) | 1.30 (0.97,1.64) | 1.68 (0.57,2.79) | 1.76 (0.41,3.12) |
| **Work** | 0.90 (0.71,1.10) | 1.09 (0.71,1.47) | 0.94 (0.75,1.13) | 0.86 (0.61,1.11) | 1.22 (0.89,1.56) | 1.43 (0.32,2.54) | 1.69 (0.34,3.05) |
| **Others** | 0.62 (0.42,0.81) | 1.10 (0.71,1.48) | 0.59 (0.40,0.78) | 0.81 (0.56,1.06) | 1.41 (1.07,1.75) | 1.87 (0.76,2.98) | 1.74 (0.39,3.10) |
|  | CH | FI | LT | GR | SI |  |  |
| **All** | 5.52 (4.78,6.27) | 5.32 (4.47,6.17) | 6.28 (4.95,7.60) | 5.29 (2.36,8.23) | 6.23 (4.81,7.64) |  |  |
| **Home** | 1.31 (0.56,2.05) | 1.01 (0.16,1.87) | 1.30 (-0.02,2.63) | 1.63 (-1.30,4.57) | 1.87 (0.45,3.29) |  |  |
| **Work** | 2.34 (1.60,3.09) | 2.25 (1.40,3.10) | 2.21 (0.89,3.54) | 2.58 (-0.36,5.51) | 2.62 (1.20,4.04) |  |  |
| **Others** | 1.88 (1.13,2.63) | 2.06 (1.21,2.91) | 2.77 (1.45,4.09) | 1.10 (-1.84,4.03) | 1.75 (0.33,3.16) |  |  |
|  | EE | HU | SK | MT | HR |  |  |
| **All** | 6.19 (5.53,6.84) | 4.48 (3.61,5.36) | 5.66 (4.96,6.35) | 8.31 (7.21,9.41) | 6.32 (5.86,6.77) |  |  |
| **Home** | 1.35 (0.69,2.00) | 1.58 (0.71,2.46) | 1.62 (0.93,2.32) | 2.19 (1.09,3.29) | 1.93 (1.47,2.38) |  |  |
| **Work** | 2.46 (1.81,3.12) | 1.75 (0.87,2.62) | 2.09 (1.40,2.79) | 2.59 (1.49,3.69) | 2.28 (1.82,2.74) |  |  |
| **Others** | 2.39 (1.73,3.04) | 1.17 (0.29,2.04) | 1.96 (1.27,2.65) | 3.54 (2.44,4.64) | 2.12 (1.67,2.58) |  |  |

**Supplementary Material IV**

<u>supp_table_forest.csv</u>

